# Nano Sensing for Early Diagnosis of Pancreatic Cancer

**DOI:** 10.1101/2023.02.21.23286244

**Authors:** Sidarth Krishna, Arthur McClelland, Tingying Helen Zeng

## Abstract

Pancreatic Cancer is becoming the second leading cause of cancer deaths, mainly attributed to late diagnosis. Surgical resection remains the only plausible treatment for curing patients of this cancer, but this is only possible when the cancer is localized making early detection vital. Currently, the most common early diagnostic method is the tri-phasic pancreatic-protocol CT scan. This method however has a low accuracy and specificity making it an ineffective early diagnostic. This purpose of this research was to develop a non-invasive, fast, and highly sensitive early diagnostic method for pancreatic cancer through the applications of nanotechnology. Surface Enhanced Raman Spectroscopy (SERS) is an innovative nano sensing method which results from plasmonic effect through molecules interacting with the surface of metal nanoparticles. This is a new method for trace biomarker detections and shows great potential to be an early nano-diagnostic method for Pancreatic cancer. This project uses Leucine as a biomarker for the demonstration of SERS for early pancreatic cancer detection. This is because recent studies show that Leucine has linked its overexpression with pancreatic cancer; studies have found a specificity of 100% with the use of Leucine and early diagnosis 2 to 5 years earlier than current diagnostic methods for pancreatic cancer. The parameters for SERS enhancement were optimized for the detection of Leucine using 20 nm Ag NPs. Predictive curves with linear fits were generated from an analysis of feature peaks giving the highest sensitivity for trace concentrations. This new finding shows the promising development of an early diagnostic method that is non-invasive, efficient and highly accurate for pancreatic cancer through SERS nano sensing.

## I. Introduction

Pancreatic Cancer is currently the fourth leading cause of cancer related deaths worldwide [2]. This form of cancer has been growing steadily and is projected to become the second leading cause of cancer deaths over the next decade [1]. Although pancreatic cancer only makes up about 3 percent of all cancer cases, it is deadly, accounting for more than twice the percent of cancer deaths at 7 percent [3]. In the US alone, more than 60,000 individuals are diagnosed yearly with pancreatic cancer while almost 50,000 will die [3]. Although there have been many developments for advancements and breakthroughs in other forms of cancer, pancreatic cancer has not improved along with this trend.

The most common and deadly form of pancreatic cancer is pancreatic adenocarcinoma (PDA) at over 85 percent of cases [1]. Pancreatic cancer results from mutations in the DNA in the pancreas, an organ located inside the lower stomach of the body [4]. This form of pancreatic cancer, PDA, begins in the cells in the lining of the ducts of the pancreas that produce digestive enzymes [4]. Common risk factors include smoking, family history, high fat diet and obesity [2]; however, treating and diagnosing pancreatic cancer remains very challenging.

Current treatments for pancreatic cancer include chemotherapy, radiation therapy and most commonly surgery [3]. Of these major treatment methods, only surgical resection of the pancreas can cure patients of this lethal cancer. For surgery to be used, the pancreatic cancer needs to be in a localized state where resection of the pancreas is possible [2]. In order to catch pancreatic cancer in these early stages, early detection is vital.

Even with the technological advancements and breakthroughs with other forms of cancer diagnosis and treatments, pancreatic cancer has not improved along with this trend [1]. There have been many studies that have tried to determine a method for the early diagnosis of pancreatic cancer, but due to multiple characteristics of pancreatic cancer this poses a large challenge.

Firstly, symptoms of pancreatic cancer include weight loss, abdominal pain, loss of appetite and itchy skin [4]. These symptoms are not observable in early stages of the cancer and rather are developed in the advanced stages of pancreatic cancer when it has spread throughout the body [4]. These characteristics result in pancreatic cancer being very difficult to effectively diagnosis based off symptoms during its early stages.

Currently, the most common diagnostic tool for pancreatic cancer is a tri-phasic pancreatic-protocol CT scan [2]. This method utilizes x-rays to create a three-dimensional visualization of the pancreas providing an accuracy of 83.3%, specificity of 43% and sensitivity of 81.4% [2] [19]. Other techniques and instruments that have been experimented with for early detection include MRI and endoscopy ultrasound [2].

These techniques are ineffective diagnostic tools as they do not have the level of accuracy, sensitivity and specificity to be an efficient early diagnostic method—typically each of these values are greater than 90% in an initial screening test. Furthermore, there are many other obstacles for these tools to become widely used. Firstly, these tools are used for patients who appear to have a family history of pancreatic cancer [2]. Studies have highlighted, however, that this segment of patients only make up about 7-10 percent of all pancreatic cancer resulting in more than 90 percent of individuals not being diagnosed until advanced stages of pancreatic cancer [2]. Furthermore, each of these techniques are time consuming, expensive and require advanced operators and researchers in order to analyze and diagnosis the information.

As a result, more than 80 percent of patients are diagnosed with pancreatic cancer at a stage that is too advanced for surgical treatment, resulting in a 5-year survival rate of under 5 percent [9]. This is much lower than many other common forms of cancer including over 90 percent for breast cancer and 88 percent for prostate cancer [6]. In order to improve the survival rate of this cancer a more effective early diagnostic method must be used in order to cure and save more patients.

For an ideal early diagnostic test for pancreatic cancer, it should be fast, efficient, inexpensive while being applicable to the entire population and most importantly with a high accuracy [1]. Many other forms of cancer diagnoses utilize multiple biomarkers in the blood and human body as an initial screening test [1]. These biomarkers become overexpressed months, or even years before symptoms emerge, making it very effective in diagnosing specific cancers [11].

Carbohydrate antigen (CA19-9) is the currently a biomarker used by the medical industry for pancreatic cancer [1] [5]. Studies have shown that the sensitivity and specificity on pancreatic cancer patients with CA19-9 was only 79% and 80% respectively [7]. These rates are not one of an effective biomarker due to the large room for error and is therefore not targeted towards the entire population worldwide as an initial screening test [1].

This research study focuses on recent advances in serum biomarkers for pancreatic cancer in order to develop a more effective and efficient early diagnostic screening test. From the analysis of multiple research studies, leucine, one of the 20 amino acids in the body, has gained attraction as a new biomarker for pancreatic cancer. It is both highly specific with optimal findings from experimental studies of 100% with diagnosis 2 to 5 years in advance of current methods [8] [10] [11].

However, to our knowledge there are no effective early diagnostic treatments for pancreatic cancer, due to the challenges in determining the expression of leucine in the blood stream. The search for a highly effective and sensitive technique to measure leucine levels in the blood is imperative to create this diagnostic method.

Surface enhanced Raman Spectroscopy (SERS) using metal nanoparticles is a new method that can be utilized as an analytical technique that is highly specific and accurate [13]. This technique has gone through rapid developments in its applications for cancer diagnosis and treatments [14]. SERS utilizes the Raman (inelastic) scattering of molecules to achieve plasmonic effect for the determination of chemical identity with vibrational modes and molecular bonds [14]. Furthermore, Raman scattering provides an efficient, accurate and noninvasive method for determining the fingerprint (chemical makeup) of molecules in any type of solution [13]. There have been recent developments of Raman scattering devices to make it portable and handheld as well providing its usefulness in any circumstance [14].

## II. Materials and Methods

### A. L-Leucine

Leucine is one of the essential amino acids in the body. Leucine C6H13NO2, a aliphatic amino acid, is vital for the synthesis of proteins in the body while also being essential for tissue regeneration and metabolism [16]. Leucine is part of the Branched Chain Amino Acids (BCAAs) which are metabolites in the body. Over the past decade, leucine has undergone multiple studies linking it to pancreatic cancer diagnosis, making it into a new potential biomarker molecule. A research study highlighted, using a transgenic PDC rat-model, the presence and expression of LRG-1 in rat pancreata developing pancreatic cancer [8]. LRG-1, leucine-rich α2-glycoprotein-1, is a serum protein in the blood stream with over eight leucine repeats and an optimal specificity of 100 percent for pancreatic cancer [8]. Furthermore, other studies have highlighted the significance of leucine expression as a BCAA 2 to 5 years in advance to developing pancreatic cancer [10]. Experimental studies have highlighted those elevated levels of leucine in the body result in a 2 times higher likelihood of developing pancreatic cancer [10]. This increase in leucine concentration have been driven by the *Kras* expression, where there is a breakdown of tissue proteins including leucine in the body as an early-stage buildup for pancreatic cancer [10].

Due to the research surrounding leucine and its developments as a serum protein marker for pancreatic cancer in early diagnostic it was chosen to be studied in depth in this research study. Specifically, the L-leucine configuration was chosen as it is the most common isomeric form of the leucine amino acid in the body [16].

### B. Surface Enhanced Raman Spectroscopy

Raman Spectroscopy has had a large increase in applicability to many fields over the last four decades [14]. It utilizes Raman scattering which is the inelastic collisions of molecules resulting in a reflective ray different from the incident ray. Using this phenomena Raman Spectroscopy was developed to produce Raman scattering for different molecules producing a unique chemical signature for each molecule based on specific chemical makeup and bond type [13]. Modifications have been studied to enhance the results and the Raman effect even more. One of these techniques is Surface Enhanced Raman Spectroscopy which utilizes metal nanoparticles, typically Ag or Au, to concentrate electromagnetic energy through surface plasmons [13]. This allows the biomarker and molecule of interest to form on the “tips” of the metal nanoparticles creating a highly localized electromagnetic field which thereby increases the magnitude of the Raman scattering effect in each molecule [13]. This makes SERS into a very effective tool for determining trace concentrations. SERS has gained traction in the medical industry for cancer diagnosis with the ability to determine chemical signatures through a noninvasive method [14]. It has performed with sensitivities and specificities of greater than 95 percent for diagnosing multiple forms of cancer including stomach, gastrointestinal and more [14]. SERS has also provided the ability to determine very small concentrations of molecules in solution even on the micro and nano scale [13]. Thus, in our study we examined the usage of Raman Spectroscopy, and specifically SERS for creating an effective, efficient and highly accurate early diagnostic tool for pancreatic cancer.

### C. Experimental Procedures

To perform the experiment four different groups were designed. The first group consisted of the control with the L-leucine dissolved in distilled water at multiple concentrations. This group was used to determine the signature Raman peaks of l-leucine to easily determine the presence of leucine amongst other chemicals for trace diagnosis.

For the second part of the experiment L-leucine solutions were mixed separately with 20 nm gold nanoparticles and 20 nm silver nanoparticles at the different concentrations. This part of the experiment was designed to demonstrate the Surface Enhanced Raman Spectroscopy (SERS) effect on the l-leucine molecule. Secondly, a 1 sample T-test will be performed. This will be used to determine the significance of Raman feature peak enhancements with NPs and the most effective nanoparticle (Au or Ag) for trace diagnosis.

The next part of the experiment is to study the size effect of nanoparticles on SERS with the L-leucine standards mixed and analyzed with a 60 nm nanoparticle size.

Lastly, the fourth group included a synthetic blood demonstration, with the leucine spiked into the samples at the same concentrations. To perform this demonstration with the blood groups one, two and three were repeated with the use of synthetic blood instead of water to create the solutions. Synthetic blood was used to represent a human blood sample at multiple stages of pancreatic cancer with leucine, while varying the SERS effect with nanoparticle size for the most effective Raman spectra.

For each experimental group, specific feature peaks on the Raman spectra can be selected and analyzed to determine a trace diagnosis of L-Leucine in solution.

### D. Sample Prepartion of L-Leucine

To create the first group of samples of l-leucine, a serial dilution was used. Studies have highlighted that the l-leucine concentration in the body typically varies around 80 to 320 μmol/L [15]. In order to represent this range of concentrations the serial dilution was specially designed to represent concentrations higher and lower than this interval for a wide range of results for the Raman spectroscopy. Furthermore, these concentrations were created to represent Pancreatic patients at multiple stages with different trace concentrations of L-Leucine in the blood stream.

L-leucine was purchased from Sigma-Aldrich and stored at 5ºC. The first concentration created was a 0.01024 mol/L solution Using a serial dilution factor of ½, 10 concentrations of L-leucine were created from a range of 20 μmol/L to 0.01024M including concentrations at both 80, 160 and 320 μmol/L to represent individuals at normal L-Leucine concentrations. Aluminum foil pieces were cut 1 cm by 1 cm to be the substrate for the Raman Spectroscopy.

Using a micropipette, 3 drops of each concentration were placed onto each piece of aluminum foil. Samples were each dried overnight forming a thin layer.

To create the samples with the nanoparticles, each of the specific nanoparticle sizes 20 nm Au, 20 nm Ag and 60 nm Ag were mixed separately with each of the L-leucine standards and micro pipetted onto aluminum foil pieces.

For the synthetic blood demonstration, the serial dilution with the l-leucine was recreated, dissolving the l-leucine in blood instead of water. The nanoparticle solutions were mixed with the synthetic blood, and each were pipetted onto separate aluminum foil pieces after being dried.

### E. Data Collection

After each of the samples dried overnight, Raman spectrum data for all the samples were collected using the Horiba LabRAM HR Evolution, a confocal Raman microscopy SERS Raman spectrum were generated. Samples used a 532 nm green laser, with a 10% laser and acquisition times of 5 seconds or 10 seconds for the shutter imaging. These specific measurements were used to maximize the Raman scattering effect with sufficient energy to generate the most effective Raman spectra for each solution with minimal fluorescence. Initial calibration was performed using a silicon wafer for the laser and diffraction grating for generating the Raman spectrum. The Raman data was averaged together from three different spots chosen on the dried layer of the L-Leucine samples.

### F. Data Processing

Once each of the Raman Spectrum were collected a data analysis was performed to identify the major peaks. In most Raman spectrum there is fluorescence emission which results when the excitation laser does not provide sufficient energy. As a result, fluorescence provides noise to Raman peaks which results in decreasing the signal and effectiveness of Raman features. To avoid the effect of fluorescence on the Raman spectrum, multiple Raman spectra were generated and analyzed for each solution. Firstly, through a background elimination process, points along the spectra were generated using a linear least squares line fit selected with a degree of 20 and max points of over 150. Points along this line were then subtracted from the spectra keeping only the Raman peaks of the l-leucine. Cosmic rays were also removed from the spectra to keep only important information in the spectra. To account for any acquisition time variability between samples the values were each scaled together to keep each of the Raman peaks constant and easy to compare to each other. For each standard, Raman spectra were averaged together to remove variability. The resulting spectra was then used for further analysis.

## III. Results

### A. Expression of L-Leucine in Raman Spectra

L-leucine samples from concentrations ranging from 20 μmol/L to 0.01024M were recognized with Raman spectroscopy. Ten samples were generated from a serial dilution with a factor of ½. Three Raman spectra were taken for each sample and averaged together to remove variability in the data. The Raman Spectra of the 0.01024 mol/L standard is shown in Figure 1. The corresponding bond type to each Raman peak at the experimentally gathered wavenumber(cm^−1^) is shown in Table II for the L-Leucine [18]. Image taken from the confocal Raman microscope of L-Leucine crystals at a concentration of 64 μmol/L is shown in Figure 2.

**Fig. 1.**
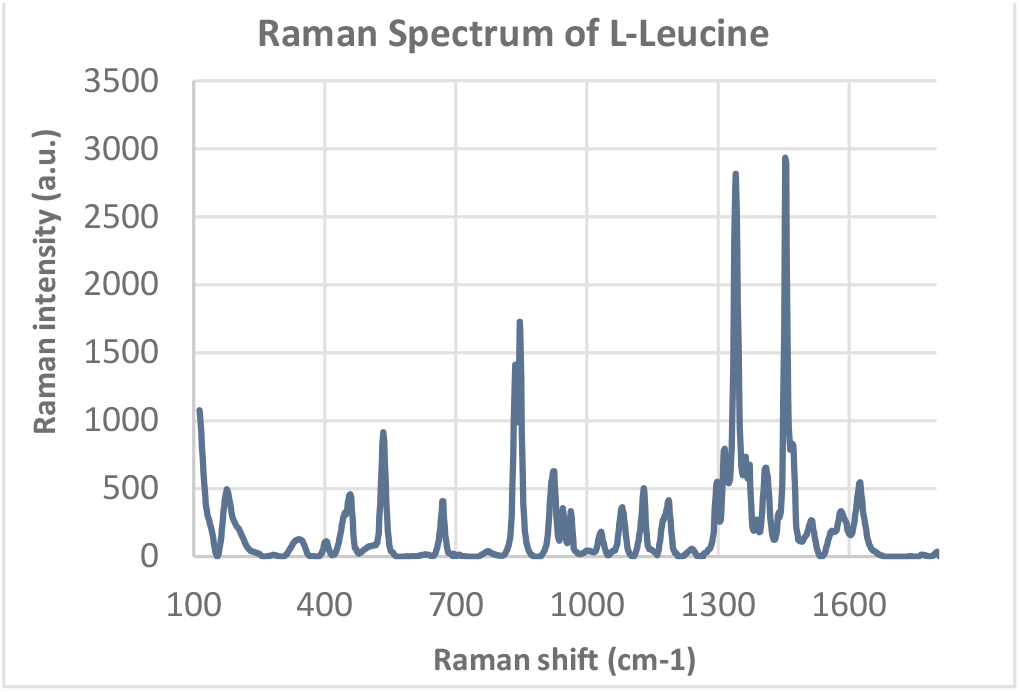
Raman spectrum at 525 nm from wavenumbers 100 cm-1 to 1800 cm-1 of the 0.01024 mol/L L-Leucine solution dissolved in distilled water. Peaks highlight the functional groups and bond types of the L-leucine molecule at different wavenumbers shown in Table II.

**TABLE I.** Experimental Design Groups

| Group Type | Components in Sample |
| --- | --- |
| Control Sample | L-leucine dissolved in distilled water |
| Nanoparticle Type | 20 nm Au NP with L-leucine water solution<br>20 nm Ag NP with L-leucine water solution |
| Nanoparticle Size | 60 nm Ag NP with L-leucine water solution |
| Synthetic Blood Demonstration | L-leucine dissolved in synthetic blood |
|  | 20 nm Ag NP with L-leucine blood solution |
|  | 60 nm Ag NP with L-leucine blood solution |

**TABLE II.**
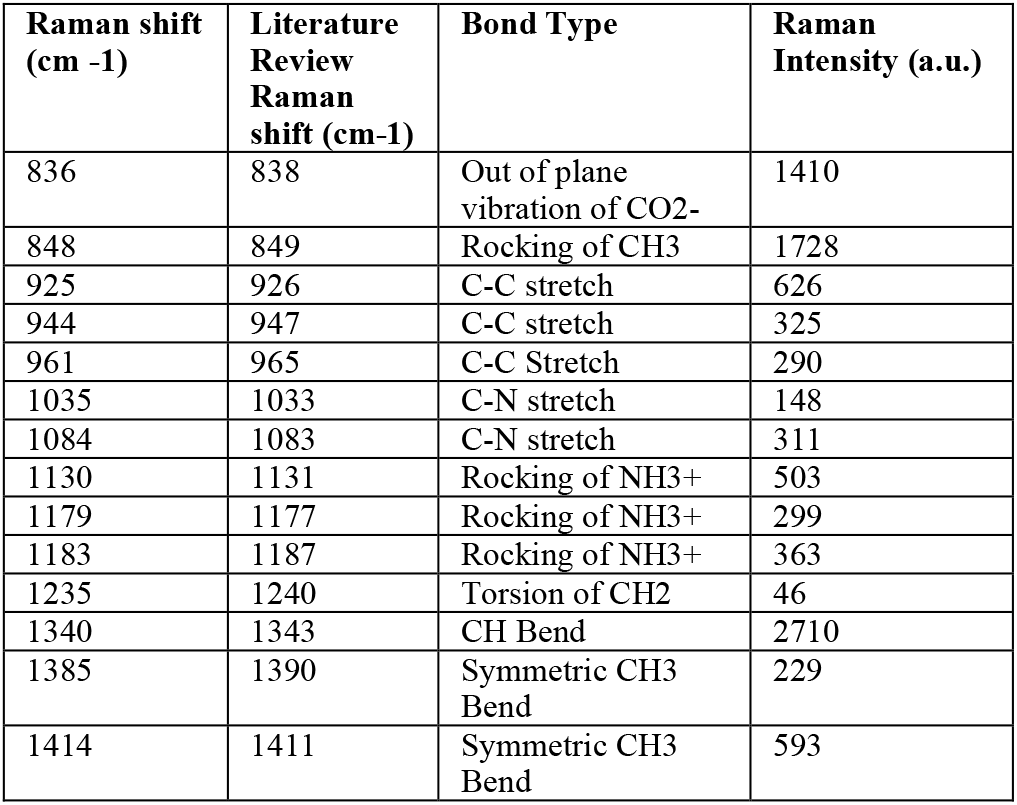
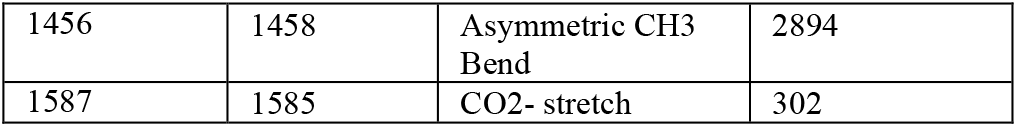
Raman Feature Peaks for L-Leucine

**Fig. 2.**
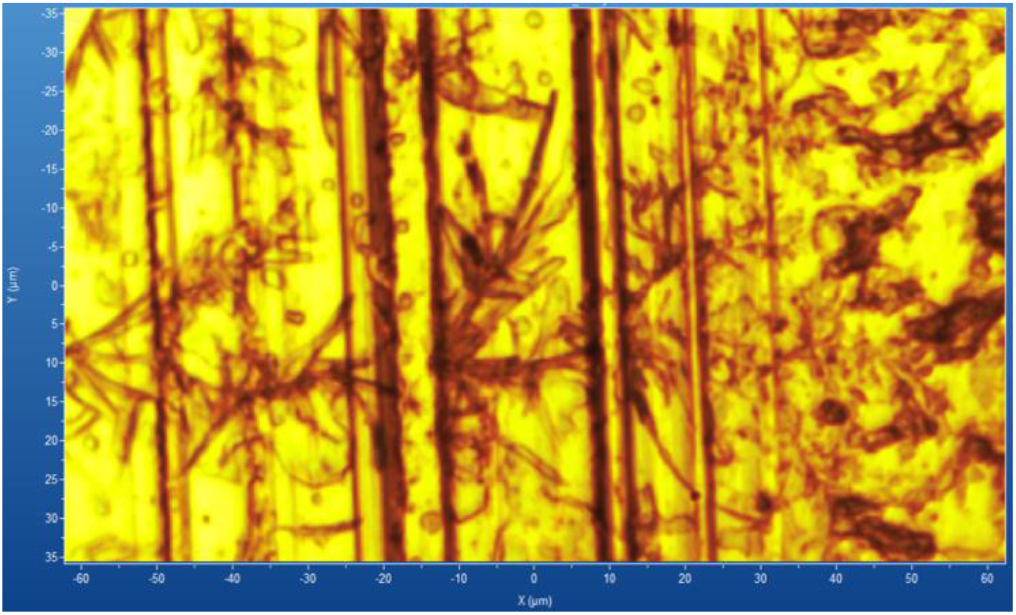
Image taken of L-Leucine crystals with the Raman confocal microscope of the 64 μmol/L L-Leucine Solution. Dimensions of image are 120 μm x 70 μm. Colors created to highlight the structure of the L-Leucine crystals. Pictured in red are the L-Leucine crystals and yellow is the aluminum substrate.

### B. Demonstration of SERS effect

Each of the 10 concentrations of L-leucine were prepared separately with 20 nm Ag Nanoparticles, 20 nm Au Nanoparticles and no nanoparticles. Three Raman spectra were generated at each concentration with the Ag NPs, Au NPs and no NPs and results were averaged together to remove variability. Figure 3 shows the Raman spectrum of L-Leucine, L-Leucine with 20 nm Ag NP and L-Leucine with 20 nm Au NP at a fixed concentration (0.01024 M). Qualitatively L-Leucine with Ag NPs has the highest Raman intensity at feature peaks, while L-Leucine with Au NPs has the lowest.

**Fig. 3.**
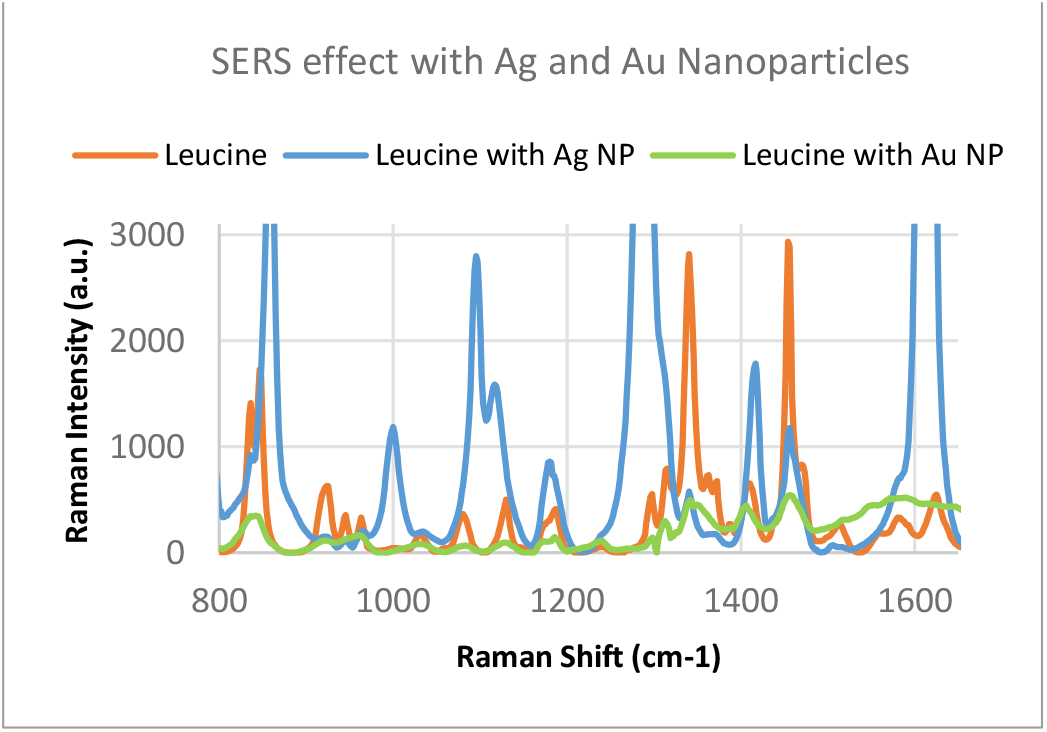
Raman spectra at 525 nm of the L-Leucine solution at 0.01024 M from wavenumbers 800 to 1650 cm-1. Group chosen to highlight the SERS enhancement of the 20 nm Ag NP and 20 nm Au NP with the Leucine.

### C. Quantification of SERS effect on L-Leucine Feature Peak

The SERS effect was quantified based on the enhancement (%) of the Raman intensity (a.u.) at feature peaks of the L-Leucine molecule. Enhancement degree was calculated as the difference between Raman intensity with Ag/Au NPs and no NP divided by Raman Intensity with no NP. All peaks except for outliers were used to calculate the average Raman peak surface enhancement degree as shown in Table III. For Ag NP this was 118.95%. The average Raman peak surface enhancement degree with the Au NP was −53.02%. On average the Ag NPs provided a 171.96% greater enhancement degree of significant Raman feature peaks than Au NPs. The surface enhancement of the Ag NPs was statistically significant (P-value = 0.041). In our study, the surface enhancement of the Au NPs was statistically significant in providing negative enhancement in the Raman Spectra of the L-Leucine (P-value = 6.425×10^^^−7). Further continued study about Au NPs is necessary for conclusions. This paper focuses on Ag NPs.

**TABLE III.** SERS Enhancement Degree (%) of Ag and Au Nanoparticles

| Raman Shift (cm-1) | Bond Type | 20 nm Ag NP Enhancement Degree (%) | 20 nm Au NP Enhancement Degree (%) |
| --- | --- | --- | --- |
| 836 | Out of plane vibration of CO<sub>2</sub>- | -35.53 | -75.82 |
| 848 | Rocking of CH<sub>3</sub> | 157.69 | -82.00 |
| 944 | C-C stretch | 63.69 | -62.15 |
| 961 | C-C Stretch | -32.06 | -52.75 |
| 1035 | C-N stretch | 33.10 | -50.00 |
| 1084 | C-N stretch | 780.70 | -84.21 |
| 1130 | Rocking of NH<sub>3</sub><sup>+</sup> | 209.34 | -85.48 |
| 1179 | Rocking of NH<sub>3</sub><sup>+</sup> | 144.81 | -64.88 |
| 1183 | Rocking of NH<sub>3</sub><sup>+</sup> | 87.05 | -69.42 |
| 1385 | Symmetric CH<sub>3</sub> Bend | -47.59 | -3.93 |
| 1414 | Symmetric CH<sub>3</sub> Bend | 200.84 | -47.04 |
| 1456 | Asymmetric CH<sub>3</sub> Bend | -62.85 | -82.41 |
| 1587 | CO<sub>2</sub>- stretch | 174.50 | 70.86 |
| Average |  | 118.94 | -53.02 |
| Standard Deviation |  | 224.9562762 | 43.34442499 |
| P-value of Enhancement Significance (1 tailed T test) | | 0.0404 (Enhances) | $6.425 \times 10^{-7}$ (Does not enhance) |

### D. Nanoparticle Size versus SERS Enhancement

The nanoparticle size of the Ag NP and its effect on the SERS feature peak enhancement was studied to determine the optimal parameters of Raman Spectroscopy for L-Leucine trace concentrations. Two particle sizes: 20 nm Ag and 60 nm Ag were studied. This comparison was run with a synthetic blood demonstration at a L-leucine concentration level of 160 μM to represent a clinically relevant concentration of the average healthy patient without pancreatic cancer. The resulting Raman spectra between the 20 nm and 60 nm Ag NP with L-leucine in synthetic blood at 160 μM is shown in Figure 4. Every feature peak of the L-leucine with 60 nm Ag NP provided a larger Raman peak intensity enhancement. The SERS enhancement of the 60 nm Ag NP in Synthetic Blood over the 20 nm Ag NP for L-Leucine is statistically significant (P-value < 0.05).

**Fig. 4.**
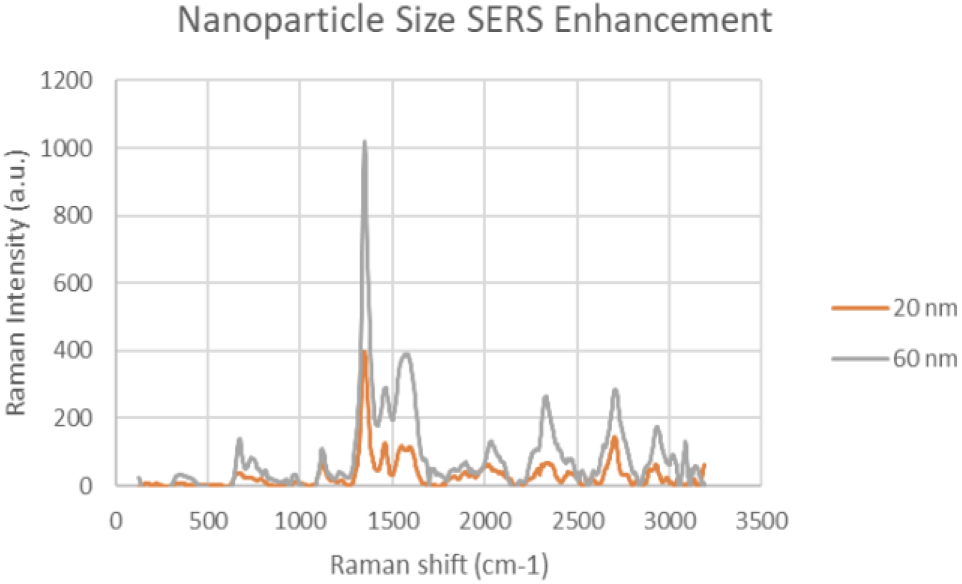
Raman spectra of 20 nm Ag NP and 60 nm Ag NP in Synthetic Blood with L-Leucine concentration of 160 μM. Group chosen to highlight the SERS enhancement of the 20 nm Ag NP and 60 nm Ag NP at a clinically relevant concentration of 160 μM in Synthetic Blood.

### E. Feature Peaks of L-Leucine Standards with Ag NP

Due to the high enhancement with Ag NP, Ag NP was used for all future experimentation. Raman spectra were taken at 6 different concentrations with Ag NP and at each concentration three Raman spectra trials were averaged for results. Feature peaks were narrowed down based on previous Ag NP enhancement. Five peaks were selected including 1084cm-1 chosen for C-N stretch, 1130, 1179 and 1883 cm-1 all chosen for rocking of NH3+ and peak at 1414 cm-1for symmetric CH3 bend. For each peak, the Raman intensity for the 6 concentrations were graphed and the R^2 for the linear fit was determined as shown in Table IV. Out of the five peaks, 3 peaks had the fits with the least variance. These were at 1130 cm-1, 1179cm-1 and 1183 cm-1 and shown in figures 5,6 and 7. The raw Raman spectra at 1130 cm-1 and the 1179cm-1/1183cm-1 are shown in figures 8 and 9 respectively.

**TABLE IV.** Linear Fitting of Raman Feature Peaks for Trace Diagnosis

| Concentration (M) | Wavenumber (cm-1) of Feature Peak |  |  |  |  |
| --- | --- | --- | --- | --- | --- |
|  | 1084 | 1130 | 1179 | 1183 | 1414 |
|  | Raman Intensity (a.u.) |  |  |  |  |
| 0.01024 | 2739 | 1556 | 861 | 727 | 1143 |
| 0.00512 | 129 | 419 | 203 | 197 | 827 |
| 0.00256 | 113 | 162 | 93 | 113 | 469 |
| 0.00128 | 82 | 161 | 76 | 62 | 721 |
| 0.00064 | 124 | 140 | 90 | 98 | 614 |
| 0.00032 | 39 | 152 | 40 | 20 | 290 |
| Linear R<sup>2</sup> | 0.8034 | 0.9062 | 0.9017 | 0.9221 | 0.7577 |
| Exponential R<sup>2</sup> | 0.9873 | 0.9972 | 0.9974 | 0.9904 | 0.7458 |

**Fig. 5.**
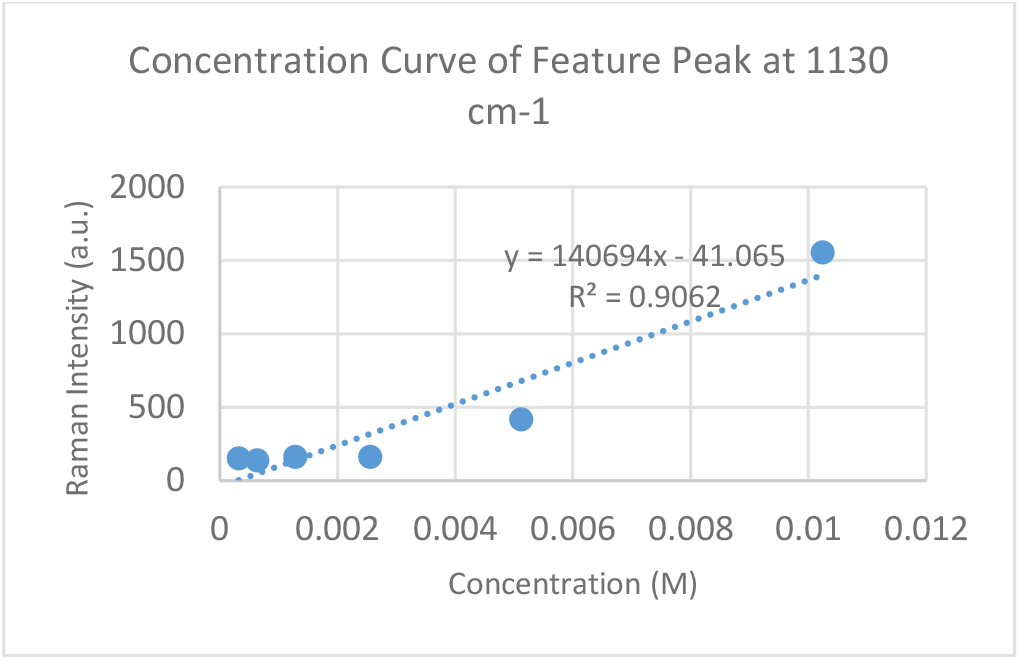
Concentration Curve generated from Raman spectroscopy data at 1130 cm-1 feature peak. The R^^^2 value is 0.9062.

**Fig. 6.**
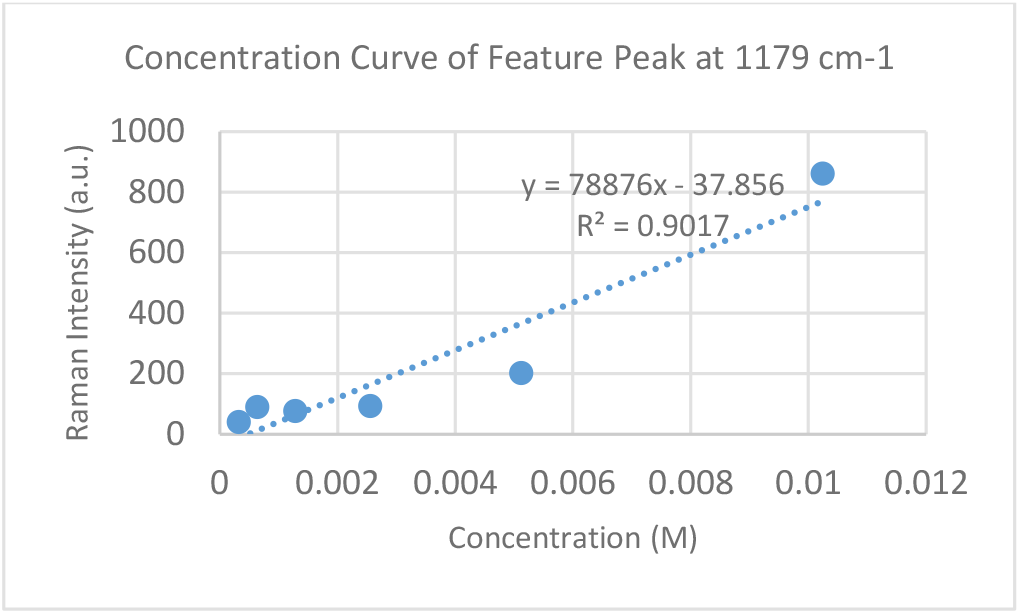
Concentration Curve generated from Raman spectroscopy data at 1179 cm-1 feature peak. The R^^^2 value is 0.9017.

**Fig. 7.**
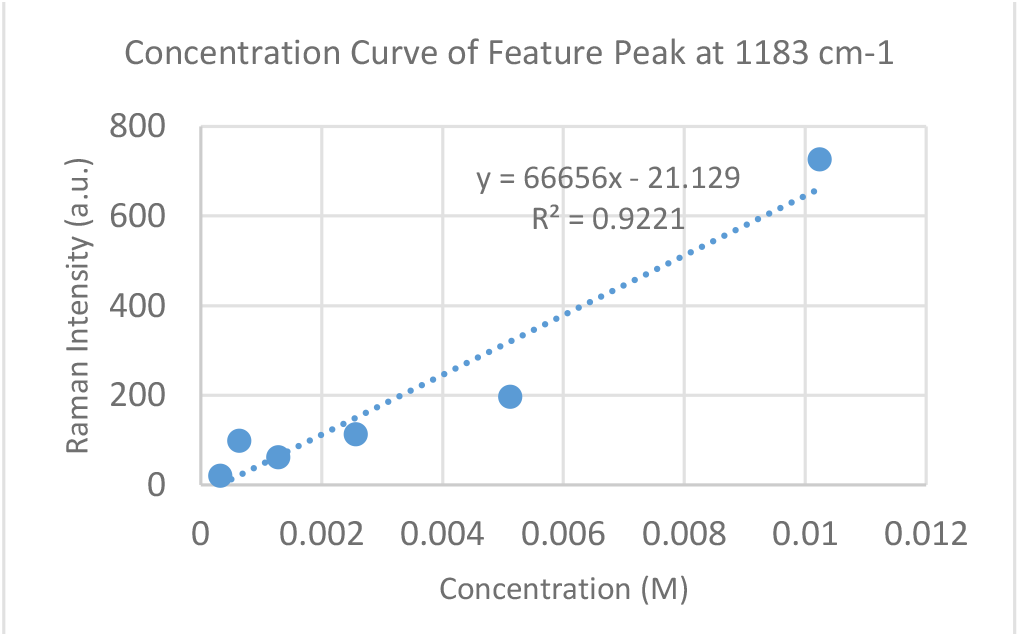
Concentration Curve generated from Raman spectroscopy data at 1183 cm-1 feature peak. The R^^^2 value is 0.9221.

**Fig. 8.**
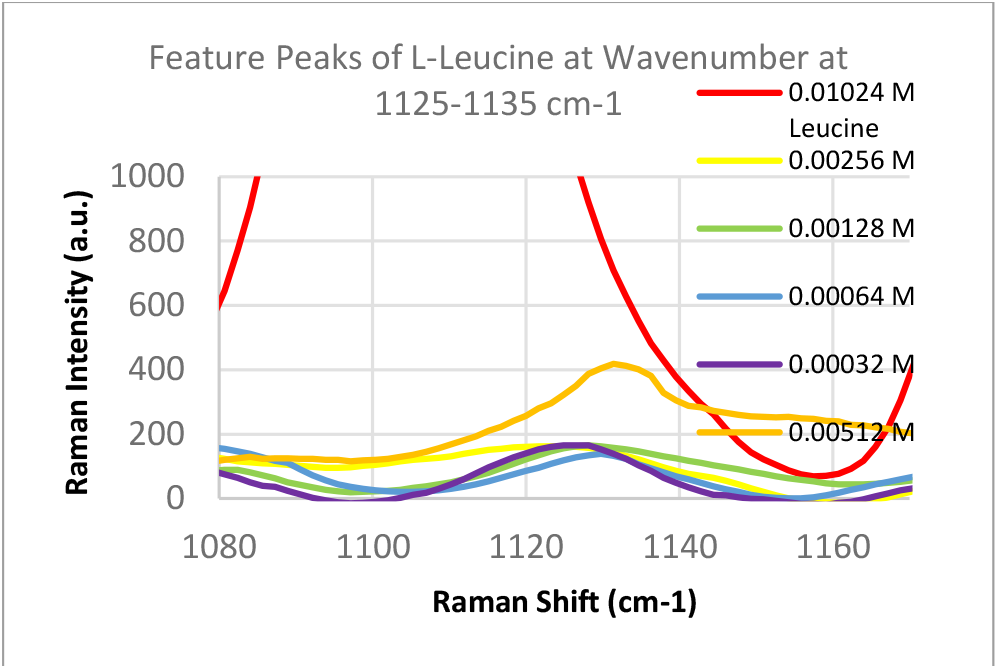
Raman spectra of L-Leucine standards from 1125 to 1135 cm-1 feature peak including feature peak at 1130 cm-1.

**Fig. 9.**
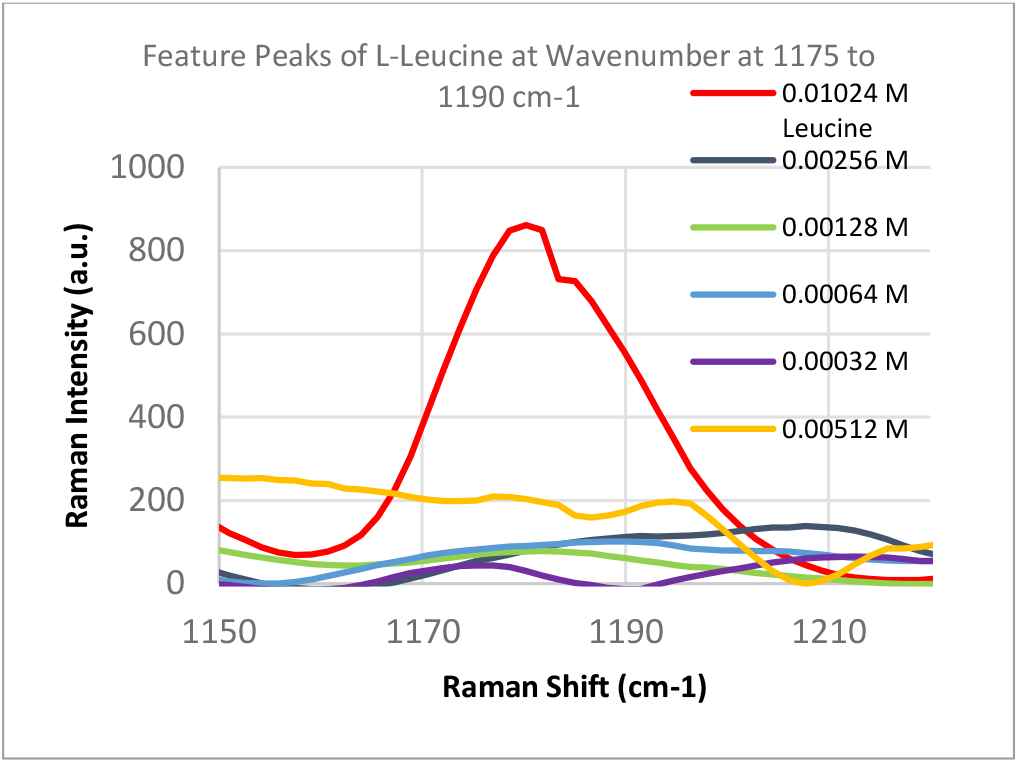
Raman spectra of L-Leucine standards from 1150 to 1220 cm-1 feature peak including both feature peaks at 1179 cm-1 and 1183 cm-1.

## IV. Discussion

In this study, an potentially early diagnostic method that was both accurate and cost-effective was developed for pancreatic cancer using L-Leucine as a biomarker. Pancreatic cancer, and specifically PDA, pancreatic adenocarcinoma is one of the deadliest forms of cancer with low survival rates and minimal treatment methods. Early diagnosis is essential, because treatments are only effective when PDA is in a localized state. In our research,, Raman Spectroscopy coupled with nanotechnology is explored as a novel, cost effective diagnostic method, called as SERS nanosensing

L-leucine has previously been shown to be a highly specific and sensitive biomarker for PDA in rodents and humans [8] [10] [11]. However, early diagnostic tests have not been developed centering around the use of L-Leucine. This is due to the challenges of detecting trace concentrations of L-Leucine in the blood stream without special instruments and tests. This paper highlights the innovative application of Raman spectroscopy in determining L-Leucine molecules in solutions lower than 20 μmol/L. Metal nanoparticles of Au and Ag were experimented with Raman Spectroscopy to create a SERS effect and test to find potential enhancements in determining trace signatures of L-Leucine. Ag NPs resulted in the largest Raman intensity enhancements with optimal peak enhancements of over 118% and a statistically significant enhancement in peak intensity (P-value < 0.041). A plausible explanation is due to strength of the Ag-O chemical bonding compared to the attraction between the Ag particles with the L-leucine. As a result, Ag NPs were used for further experimentation due to its increased sensitivity towards the detection of L-leucine. Ag NP sizes of 20 nm and 60 nm were also experimented with using a Synthetic Blood demonstration to determine the optimal nanoparticle size for the SERS enhancement of detecting L-Leucine in the blood. 60 nm Ag provided a statistically significant SERS enhancement of feature peaks over the 20 nm Ag NP (P-value < 0.05).

After the Raman Spectroscopy parameters were optimized for Surface Enhancement detection of L-Leucine, the next major goal was to determine whether concentrations of L-leucine in health patients versus patients with early PDA could be well distinguished using Raman spectroscopy.

This would be proven and distinguished by a significant linear relationship between the Raman intensity and concentration of L-Leucine for a given feature peak. The six main concentrations used ranged from 0.00032 M for healthy patients up to 0.00256 M, 0.00512 M and 0.01024 M for patients with early PDA, with these values based off of previous research [11] [15].

After assessing several feature peaks from the Raman spectra with the use of Ag NP, wavenumbers 1130, 1179 and 1183 cm-1 demonstrated the highest sensitivity to concentration changes to L-leucine. Analysis of the Raman intensity of these peaks produced a predictive curve for determining the concentration of L-Leucine in unknown samples of blood with high accuracy. Synthetic blood demonstrations were also tested to determine the accuracy and predictability of L-Leucine trace concentrations through the use of feature peaks. The results upheld prior experimental data, indicating the use of these three peaks for determining the trace concentration of L-Leucine at a high sensitivity.

## V. Conclusions

This study has developed a nanosensing based method applying SERS to create an early diagnostic method for pancreatic cancer. In this primary research, we demonstrated the optimizations of Surface Enhanced Raman Spectroscopy parameters for detecting trace concentrations of Leucine, a significant biomarker for pancreatic cancer. With the use of 60 nm Ag Nanoparticles, L-Leucine concentrations at under 20 μM/L, which is 4 times lower than average concentrations of Leucine in the blood were easily detected. Enhancements were greater than 118% for feature peaks, allowing for the feasibility and high sensitivity of SERS as a potentially early diagnostic novel technology. Our findings have determined that the most significant feature peaks of leucine include 1130, 1179 and 1183 cm-1. These peaks produced predictive curves with significant linear fits that will allow for determining the overexpression of Leucine with high accuracy and sensitivity in unknown samples of blood. The proposed approach provides a non-invasive, efficient, fast and easy with a high sensitivity and accuracy early diagnostic method for pancreatic cancer.

Continued research is to make collaborations with clinical center to try this potential early diagnostic method on clinical data from blood samples. Specifically, blood samples of healthy patients as well as cancer patients at different stages of PDA. The use of portable Raman Spectroscopy devices is already available allowing for the implementation of a Ag NP SERS early diagnostic method for pancreatic cancer in the near future.

## Data Availability

The research does not involve any clinic trails nor animal models and no human patient data used. All checmicals were from Sigma aldrich. All data are Raman spectroscopy. They are available as requests.

## Acknowledgment

S. Krishna is grateful for the intern research opportunity offered by the Academy for Advanced Research and Development (AARD). This project is supported by the scholarship of AARD for Future Scholars. Raman Spectroscopy data collection and analysis were performed in part at the Harvard University Center for Nanoscale Systems (CNS); a member of the NNCI, which is supported by the National Science Foundation under NSF award no. ECCS-2025158.

## Notes

### Competing Interest Statement

This paper was developed by the sponsorship of intern research training to students in the summer of 2022 through the financial support of scholarship to the first author, Sidarth Krishna. AARD is the sponsor. Dr. Zeng is the PI for this project.

### Funding Statement

This primary study was patially funded by the Academy for Advanced Research and Development (AARD) though scholarship of the Future Scholars from AARD for the first author.

